# Decoupling of spatial scales in breast pathology reveals fractal-like nuclear organization emergent from tissue spatial architecture

**DOI:** 10.64898/2026.05.02.26352267

**Authors:** A. Das, H. Ahammer, J. S. Prabhu, R. Bhat, M. K. Jolly

## Abstract

Quantitative biophysical signatures of nuclear spatial reorganisation across breast carcinoma progression remain insufficiently characterised. We apply two complementary fractal descriptors, Correlation dimension (*Dc*) and Minkowski dimension (*Dm*), to 4276 regions of interest across seven breast tissue subtypes from the BRACS dataset, validating observed dimensions against systematically constructed null spatial models to distinguish genuine structural organisation from geometric irregularity. All subtypes significantly exceed the complete spatial randomness baseline, confirming universal departure from random nuclear arrangement. The observed scaling is characterised as statistically monofractal within a bounded pre-fractal range. Invasive carcinoma uniquely fails to exceed the clustered null in *Dc* while simultaneously showing the weakest *Dm* null deviation, a dual convergence toward stochastic baselines consistent with the progressive removal of architectural constraints. Flat epithelial atypia exhibits a unique directional dissociation with the lowest *Dc* across all subtypes combined with high *Dm* null deviation, a co-occurrence not observed in any other subtype and geometrically consistent with decoupled nuclear spatial organisation at the centroid distribution and boundary morphology scales. Interpreted within a percolation-theoretic framework, the non-monotonic null deviation trajectory maps onto qualitative regime transitions, providing a physically grounded explanation for the observed discrimination profile across pathological transitions. These findings position fractal-like nuclear architecture as a potential descriptor for pre-malignant transitional states.

## 1. Introduction

Breast carcinoma remains the most frequently diagnosed malignancy in women and a leading contributor to cancer-related mortality worldwide [1]. In clinical practice, histopathological evaluation of tissue biopsies constitutes the diagnostic standard, typically employing the Nottingham Grading System to assess tubule formation, nuclear pleomorphism, and mitotic frequency [2]. This paradigm rests, however, on subjective visual interpretation, introducing significant intra- and inter-observer variability [3], reported diagnostic disagreement rates up to 75.3% [4], and motivating the development of objective, reproducible histomorphometry descriptors [2].

Cancer is increasingly reconceptualised not as a purely gene-centric disease, as formalised by the somatic mutation theory, but as a multiscale disruption of the structural and mechanical constraints governing the epithelial-stromal interface [5]. The Tissue Organisation Field Theory (TOFT) posits that malignancy emerges from a progressive breakdown of the physical boundary conditions that normally maintain epithelial spatial order [5–6]. Within this framework, pathological states represent distinct physical configurations of the tissue system, and progression toward malignancy constitutes a structural reorganisation at the mesoscopic scale, driven by collective nonlinear interactions between biological units and their mechanical microenvironment [5]. Nuclear morphology is a direct physical manifestation of these interactions, regulated by the mechanical coupling between cytoskeletal architecture, consisting of actin stress fibres, microtubules, and the condensation state of chromatin [5,7]. At the scale of cell populations and tissue architecture, the collective spatial arrangement of nuclei within a tissue field reflects the geometric organisation of the tissue system in which they reside, making nuclear spatial distributions a measurable proxy for the architectural consequences of tissue-level reorganisation across pathological progression. Prior fractal geometry-based histopathological research has focused extensively on nuclear chromatin texture, i.e., the internal grey-value distribution within individual nuclei [8,9], which characterises intra-nuclear organisation but does not capture spatial dependencies between nuclei across the tissue field. This approach treats nuclear features as independent observations, structurally discarding the spatial correlations between neighbouring nuclei that are significantly stronger than those between distant nuclei and that encode the collective architectural state of the tissue. The present study, therefore, shifts focus to nuclear spatial architecture, conceptually interpreted as the multiscale synthesis of geometric form and spatial topology across a tissue field, capturing the breakdown of organisational order that is characteristic of malignant progression.

Critically, this reorganisation does not proceed as a continuous structural transformation. Rather, disease progression corresponds to regime switching between distinct dominant physical processes as tissue transitions between varied states of structured aggregation, critical organisational balance, and near-randomness, leading to changes in governing spatial process at a qualitative behaviour level, not merely quantitatively [10–11]. Fractal-like scaling is predicted to emerge transiently near such transitions, when the system loses a dominant characteristic length scale [12]. In normal epithelium, cell size defines a characteristic structural scale, and fractal-like behaviour is suppressed [13]; at transitional stages, structural heterogeneity spans many scales simultaneously, and a fractal-like organisation emerges. In advanced cancers, a dominant tumour mass re-establishes a characteristic scale, and fractal-like behaviour weakens. In natural systems, this scaling exists over a bounded range, i.e., between a lower cut-off at the cellular scale and an upper cut-off imposed by tissue geometry, constituting fractal-like or pre-fractal or asymptotic fractal rather than strict mathematical fractal behaviour [14–15]. Whereas Euclidean geometry cannot describe this scale-bounded complexity, fractal geometry provides the requisite framework, with the fractal dimension (FD) serving as an index of how structural detail varies across hierarchical scales.

A critical requirement for valid interpretation of FD estimates is statistical comparison against null spatial models [16]. A computed dimension in the range 1 < *FD* < 2 is a necessary but not sufficient condition for fractal-like behaviour since any geometrically irregular boundary or non-random point pattern can trivially satisfy this criterion. Null model comparison, in which observed dimensions are tested against those from stochastic processes with known, non-fractal properties, provides the statistical evidence needed to distinguish genuine structural complexity from geometric irregularity alone [16]. Consequently, separate null models are applied to nuclear spatial distributions and nuclear boundary complexity, respectively.

To quantify nuclear spatial architecture, we employ two complementary fractal measures. The Minkowski dimension (*Dm*) is computed via morphological dilation of nuclear boundary pixels, progressively probing boundary complexity across spatial scales. Notably, the dilation-based implementation confines influence to object (background or padding) pixels, minimising contributions from image padding or background. The Correlation dimension (*Dc*) is derived from pairwise spatial correlation of nuclear centroids; the correlation integral operates on detected centroid coordinates rather than pixel arrays, rendering estimates insensitive to boundary padding. Together, *Dm* captures local morphological complexity of nuclear boundaries, while *Dc* quantifies how the nuclear population fills space and maintains spatial correlation across scales. Consequently, they are geometrically complementary descriptors of the tissue architectural state.

We hypothesize that the fractality of nuclear spatial architecture constitutes a quantitative signature of pathological progression across multiple spatial scales, and that its emergence, evolution, and breakdown across the seven-stage diagnostic spectrum encode distinct physical regimes of tissue organisation. We apply this framework to the BReAst Carcinoma Subtyping (BRACS) dataset [3], spanning Normal (N), Pathological Benign (PB), Usual Ductal Hyperplasia (UDH), Flat Epithelial Atypia (FEA), Atypical Ductal Hyperplasia (ADH), Ductal Carcinoma In Situ (DCIS), and Invasive Carcinoma (IC). We systematically validate fractal-like scaling, assess dimensional complementarity and confounding influences, compare observed dimensions against rigorously constructed null models with paired non-parametric effect-size quantification, characterise within-patient spatial heterogeneity, and benchmark the clinical utility of this architectural signature through machine learning classification across all pathological transitions.

## 2. Methods

### 2.1. Data Acquisition and Preprocessing

The study utilized the BRACS histopathology image dataset containing 4276 regions of interest (ROIs) from 368 whole slide images (WSIs) of Hematoxylin and Eosin (H&E) stained images, representing a seven-stage pathological spectrum. For a scientifically meaningful scaling-based analysis, all ROIs were confirmed to originate from the native 40× acquisition level of the Aperio image pyramid [3], yielding a uniform physical resolution of 0.25 µm/pixel across the dataset. This ensures that inter-subtype geometric variability reflects underlying biological architecture rather than differential image scaling. Additionally, for computational efficiency, raw Portable Network Graphics (PNG) files were converted to Tagged Image File Format (TIFF) format images.

### 2.2. Nuclei Segmentation and Binary Mask Verification

Nuclear instance segmentation was executed using the Stardist two-dimensional (2D) algorithm via its Fiji plugin [17], which prioritizes the identification of individual star-convex objects (e.g., cell nuclei) over conventional pixel-level classification. In contrast to semantic segmentation, which merges adjacent objects into single blobs, Stardist utilizes shape-aware regression to predict the distance of each pixel to the boundary of the parent cell, subsequently allowing effective separation of crowded or overlapping nuclei within the dense pathological landscape, providing accurate boundaries for nuclear structures.

We utilized the pre-trained versatile H&E nuclei model, which has been rigorously trained on the MoNuSeg 2018 training data and The Cancer Genome Atlas (TCGA) archive [17]. To ensure precise boundary detection and optimal centroid localization, images were normalized with the complete percentile range of 1 to 100. The segmentation was further refined by setting a probability score threshold of 0.50 to balance between false positives and true negatives, and minimize false positives, and an overlap threshold of 0.30 to manage the spatial connectivity and unique identity of individual segmented objects across the multiscale tissue architecture. The resulting nuclear masks were converted to 8-bit binary format. To ensure the integrity of the binary data, histogram-based statistical analysis (mean, minimum, maximum, standard deviation, and mode) was performed across each subtype. This verified that background pixels were strictly black (pixel value = 0) and foreground nuclei pixels were strictly white (pixel value = 255).

For the null model’s comparison study concerning *Dc*, nuclear centroid coordinates were generated for each binary mask from the Fiji platform. Images containing fewer than 20 nuclear centroids were excluded from all subsequent analyses to ensure statistically meaningful FD estimation.

### 2.3. Image Dimension Standardization

Analysis of image size distribution revealed substantial variability in width (127-5023 pixels) and height (137-6005 pixels). To standardize the spatial domain for fractal analysis and meaningful comparison, all images were padded to a uniform target dimension of 5023 × 6005 pixels following global statistical comparison across the dataset, not displaying extreme deviations. Padding was performed in Fiji by increasing the canvas size and adding black pixels symmetrically in both horizontal and vertical directions. The workflow ensured the segmented nuclei pattern remained centered within the padded image.

Nevertheless, it is noteworthy that for the null model’s study, spatial extents were restricted to the original image domain dimensions or size rather than the padded canvas, ensuring that padding introduced no systematic bias into comparisons.

### 2.4. Minkowski Dimension Optimization and Computation

To optimize the measurement parameters, we selected 70 representative images (10 from each subtype) that spanned the full range of observed image size and aspect ratios (≈ 0.23-3.52). The Minkowski dimension was computed using the ComsystanJ [18] plugin in Fiji by implementing morphological dilation with a disk kernel, where the scaling exponent governs the increase in nuclear area *A* as a function of dilation radius *r* and represented by Eq. 1 [19]:

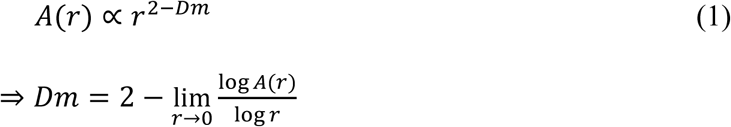

The number of dilations was varied from 3 to 20, and the qualitative trends of the mean *Dm*, *R*^2^, and standard error, respectively, were analyzed for selecting the optimized dilation range.

For nuclear boundary complexity, a geometrically meaningful null requires a smooth reference boundary preserving nuclear size and orientation while eliminating roughness; Landini and Rippin [20] demonstrated that nuclear boundaries behave as asymptotic pre-fractals exhibiting fractal-like scaling at low resolution but converging to smooth geometry at high resolution, motivating a null based on smooth matched ellipses. Image noise further inflates computed FD estimates systematically, with inflation magnitude inversely related to the space-filling character of the object [21], underscoring the need for conservative null design and statistical analyses.

To determine whether real nuclear boundaries exhibit significantly greater morphological irregularity than smooth, non-fractal boundaries of equivalent nuclear geometry, a geometry-matched smooth ellipse null model was constructed following a modified approach in consideration of ref. [20]. Here, *Dm* was re-estimated from both the original images and null realisations using a custom Python implementation of the Minkowski sausage or Minkowski-Bouligand method, in which the area *A*(*r*) of the dilation neighbourhood was evaluated via a single Euclidean Distance Transform (EDT) of the boundary image, with *A*(*r*) = #{*pixels*: *distance* ≤ *r*} obtained by vectorised thresholding across all radii simultaneously. This implementation was used to enable programmatic batch simulation and ensure that dimension estimates for real and null images were computed by an identical algorithm, thereby supporting the paired comparison.

For each nucleus in the binary mask, semi-major axis (*a*), semi-minor axis (*b*), and orientation (*θ*) were extracted from region properties. For each of 5 null realisations, ellipse parameters were independently perturbed with small Gaussian noise defined by, *a*^′^ = *a* + *N*(0,0.1*a*), *b*^′^ = *b* + *N*(0,0.1*b*), *θ*^′^ = *θ* + *N*(0,0.05*rad*), with *a*′ and *b*^′^ clipped to a minimum of 1 pixel, and each perturbed nucleus was rasterised and its boundary extracted before compositing onto the null boundary canvas. The per-nucleus extraction procedure ensures connected, closed boundary topology at every realisation, and prevents shared-edge artefacts between spatially adjacent nuclei. The mean null *Dm* across five realisations served as the per-image null estimate, and was sufficient given the narrow null distribution produced by the constrained perturbation scale. The implemented smooth ellipse null, by preserving nuclear area and orientation while eliminating boundary roughness, isolates genuine boundary complexity from noise-induced inflation.

Null realisations were imposed on each image’s nuclear geometry, i.e., centroid positions, axis lengths, orientations, and adaptive radius bounds, rendering the comparison inherently paired. Subsequently, a paired Wilcoxon signed-rank test was applied to compare real versus null *Dm* for each subtype independently. Effect size was quantified as Cohen’s d for paired differences, implementing standard deviation with Bessel’s correction.

### 2.5. Correlation Dimension Optimization and Computation

In this study, *Dc* was estimated from 8-bit binary nuclear masks using the ComsystanJ [18], implemented via the binary value mass algorithm with raster box scanning. Here, the spatial domain of the binary image is partitioned into a grid of square boxes, and the fractal measure is approximated by summing the squared count of foreground pixels within each box. It is a computationally efficient raster approximation of the Grassberger-Procaccia correlation integral [22,23]. For optimization, the box size was varied from 2 to 13, distributed according to successive powers of 2, and was used to sample the scaling relationship between box size and the spatial correlation mass and the qualitative trends of the mean *Dc*, *R*^2^, and standard error, respectively, were analyzed to select the optimized number of boxes. *Dc* was extracted as the slope of the log-log relationship between box size and the squared pixel-count sum across the scaling regime, defined by Eq. 2:

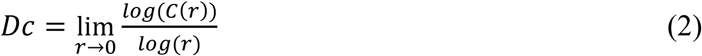

To determine whether observed *Dc* values reflect spatial organisation beyond what physically defined stochastic processes can produce, two null spatial models were applied: Complete Spatial Randomness (CSR), i.e., the Poisson process in which points or nuclei are distributed independently and uniformly, and a clustered null based on the Thomas process in which nuclei aggregate around randomly distributed parent centres [24–26]. For null model computation, *Dc* was re-estimated from both the original images and null realisations using a custom Python implementation of the Grassberger-Procaccia correlation integral [22–23], to enable programmatic batch simulation and ensure methodological consistency between real and null comparisons. For a set of *N* nuclear centroid coordinates, all pairwise Euclidean distances were computed, and the correlation integral *C*(*r*) was evaluated over 15 geometrically spaced radius values between *r*_*min*_(5th percentile of all pairwise distances) and *r*_*max*_ (25% of the smaller domain dimension) from Eq. 3 [23]:

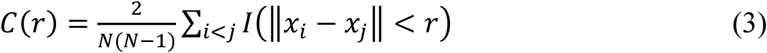

Here, ‖*x*_*i*_ − *x*_*j*_‖, and *I*(*x*) represents the Euclidean distance between centroids, and an indicator function which equals 1 when the separation between centroids is less than a threshold radius, and 0 otherwise, respectively.

In the computational implementation, self-pairs were excluded by construction, i.e., the lower radius bound at the 5^th^ percentile additionally avoids bias from spatially proximate pairs. Dc was identified as the slope of log (*C*(*r*)) versus *log*(*r*) in the power-law scaling regime, using a sliding window of width 8 log-spaced points; the window maximising *R*^2^, subject to 0 < *Dc* ≤ 2.1 was selected. As per results from ComsystanJ analysis, images for which no valid window was identified or for which the best-fit *R*^2^fell below 0.90 were excluded.

Both null models were restricted based on the observed centroid count *N* and original domain dimensions of each image, ensuring paired comparisons that control for image-level covariates. Under CSR, centroids were drawn independently from a uniform distribution over the image domain, representing the Poisson null hypothesis of no spatial dependence between nuclei [27]. Under the Thomas process, max (5, *N*/20) parent centres were placed uniformly within the domain, and *N* daughter centroids were distributed as Gaussian offsets (*σ* = 10% of the smaller domain dimension) around randomly selected parents, clipped to the domain boundary. This clustered null captures spatially structured but non-fractal aggregation, providing a stochastic baseline against which fractal-like multiscale spatial organisation can be distinguished from elementary clustering tendency. For each image, 20 independent realisations of each null model were generated, *Dc* was computed for each realisation, and the mean null *Dc* across realisations served as the per-image null estimate.

The paired Wilcoxon signed-rank test was applied to compare real versus CSR and real versus clustered *Dc* values within each subtype independently. Non-parametric testing was selected for distributional robustness and for consistency with the Kruskal-Wallis framework applied in inter-subtype comparisons. Equal numbers of real and null pairs were confirmed prior to each test.

### 2.6. Statistical Analysis

For results from ComsystanJ analysis, statistical analyses were performed to address five interrelated questions: (i) validation of fractal-like scaling behaviour; (ii) inter-subtype discrimination and pathological transition analysis; (iii) complementarity and independence of *Dm* and *Dc*; (iv) partitioning of spatial variance and within-patient heterogeneity; and (v) diagnostic classification performance. All analyses were conducted at the ROI level unless otherwise stated. Custom Python programs were used for all computations; however, qualitative trends were independently verified using GraphPad Prism 11 (trial version). Mathematical formulations and detailed statistical rationale are provided in Supplementary Material (S1).

## 3. Results

This study evaluates fractal-like nuclear spatial architecture as a biophysical signature of breast carcinoma progression. Applying principles of statistical mechanics and fractal geometry to the BRACS dataset, we quantified tissue architectural organisation across seven subtypes. Following parameter optimisation, *Dm* was computed using 10 dilation steps, which preserved a sufficiently high scaling range, high *R*^2^, moderate standard error, and minimal edge effects. *Dc* was computed using 11 box sizes, which maintained high log-log linearity, low uncertainty, and minimal sensitivity to background and finite-size effects.

### 3.1. Validation of fractal-like scaling

The presence of fractal-like scaling in nuclear architecture was systematically validated through the coefficient of determination (*R*^2^) for both descriptors. The correlation dimension (*Dc*) demonstrated fractal-like behavior with a mean *R*^2^ = 0.997 where 95.8% of all ROIs exhibiting *R*^2^ ≥ 0.99. The mean *Dc* across the subtypes was found to be 1.615 ± 0.076 (Table S1). Similarly, the Minkowski dimension (*Dm*) exhibited a mean *R*^2^ = 0.9699 with 99.3% of all ROIs showing *R*^2^ ≥ 0.95 (Fig. 1(a)). The mean *Dm* across the subtypes was found to be 1.63 ± 0.042. A quantitative comparison confirmed that the power-law scaling of *Dc*, interpreted as a multiscale descriptor of nuclear centroid spatial correlation, fits the scaling model significantly better than *Dm*, interpreted as a descriptor of nuclear boundary morphological complexity, with *t* = 197.6, *p* < 0.001 and Cohen’s d value of 3.022 (Fig. 1(b)). In addition, a paired *t*-test revealed a significant difference in mean dimension values between the two descriptors (*t* = −15.2, *p* < 0.001), consistent with their measurement of geometrically distinct architectural properties. Nonetheless, while the fit quality was consistently high across the dataset, Kruskal-Wallis test revealed that *R*^2^ values for *Dm* varied more significantly across pathological subtypes than *Dc* (Fig. 2(a)) with the test statistic *H*(6) = 54.51 vs. 14.71. Notably, *Dc* scaling quality remained invariant relative to disease progression, as revealed from the Spearman correlation test (*ρ* = −0.005, *p* = 0.927), whereas *Dm* scaling quality exhibited a size dependence (*ɛ*^2^ = 0.134) on the pathology subtype (*ρ* = −0.276, *p* < 0.001).

**Fig. 1.**
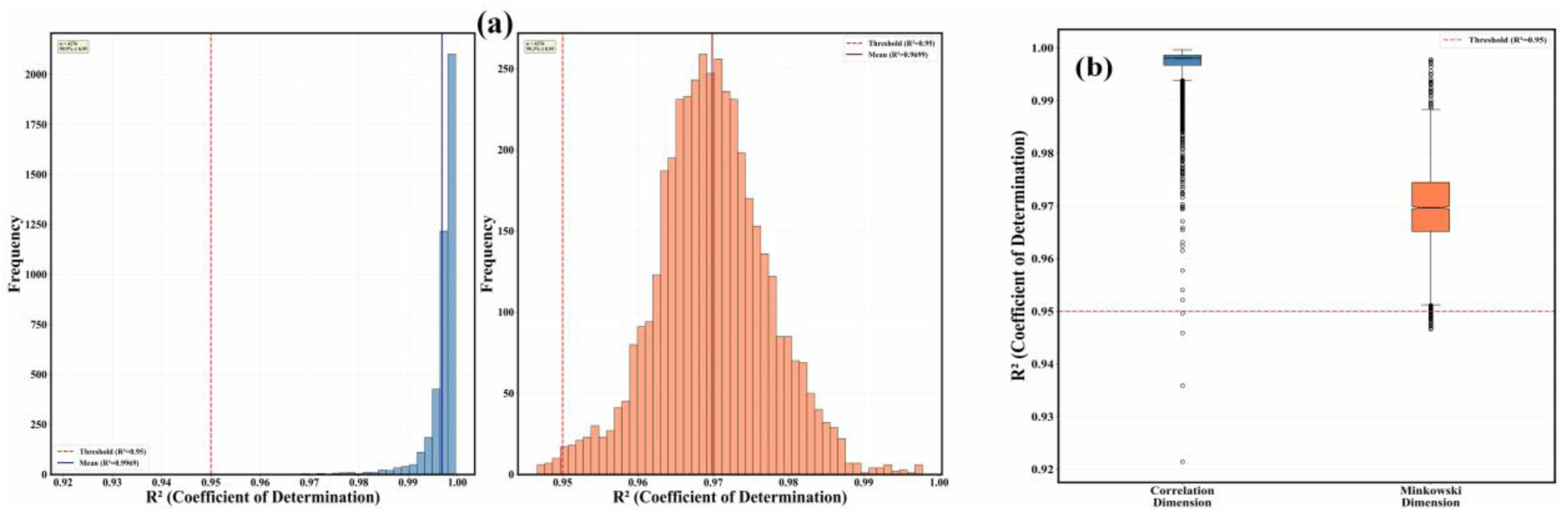
**(a)** R^2^ distribution displays excellent fits with left panel: Dc mean = 0.997 (95.8% ≥ 0.99) and right panel: Dm mean ≈ 0.976 (99.3% ≥ 0.95) and **(b)** Paired comparison confirms R^2^ (Dc) significantly exceeds R^2^ (Dm) with threshold (red-dashed line) of 0.95

**Fig. 2.**
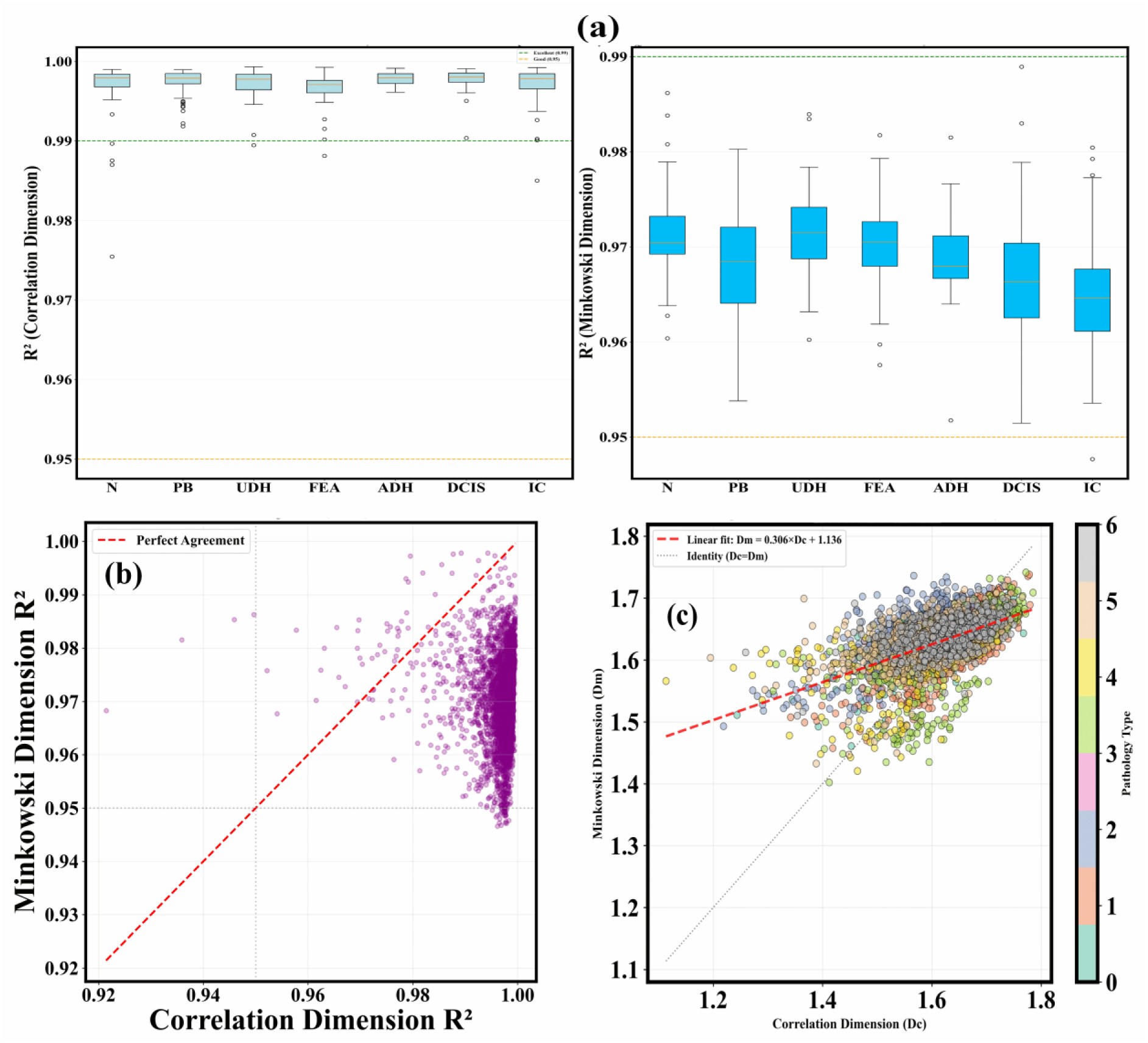
**(a)** R^2^ (Minkowski Dimension) (right panel) showing significant variation by subtypes, and declines with cancer progression, indicating scaling degradation of nuclear boundary complexity in advanced subtypes. Box plots display median (horizontal line), interquartile range (box), and outliers (circles). **(b)** Scatter plot of R^2^ (Dc) and R^2^ (Dm) exhibits moderate correlation (r^2^ = 0.31), and **(c)** Dc vs. Dm scatter, color-coded by subtype (0=N, 1=PB, 2=UDH, 3=FEA, 4=ADH, 5=DCIS, and 6=IC) demonstrates complementarity of the investigated fractal dimensions

### 3.2. Statistical validation against null spatial models

A critical requirement for interpreting FD estimates as biophysically meaningful, rather than as geometric inevitabilities of any irregular or non-random spatial pattern, is comparison against null models with known, non-fractal properties. The following results support that the observed dimensions reflect genuine structural organisation beyond stochastic baselines, and that this organisational surplus varies systematically and non-monotonically across the pathological spectrum.

All seven subtypes exhibited *Dm* values significantly exceeding those of the geometry-matched smooth ellipse null model (*p* < 10^−65^), highlighting that real nuclear boundaries carry morphological complexity not attributable to nuclear size and orientation alone (Table S2). However, the magnitude of this excess, quantified by Cohen’s *d* for paired differences, varied markedly across subtypes and did not follow the clinical progression sequence. Specifically, PB showed the largest excess (*d* = 1.475), followed by N (*d* = 1.392), FEA (*d* = 1.096), ADH (*d* = 1.012), UDH (*d* = 0.925), DCIS (*d* = 0.832), and IC (*d* = 0.423) (Table S2). IC exhibited boundary complexity only marginally above the smooth null, less than half the effect size of the next lowest group, indicating a near-collapse of morphological organisation at the nuclear boundary scale.

All seven subtypes showed *Dc* values significantly below the CSR null (*p* < 10^−35^), highlighting that no subtype exhibits spatially random nuclear distribution (Table S3). Alternatively, all tissues are more clustered than a Poisson process. The comparison with the Thomas clustered null, however, revealed a critical asymmetry. Six of seven subtypes showed *Dc* values significantly below the clustered null (*p* < 10^−7^), indicating spatial organisation beyond elementary aggregation. IC was the sole exception, with its real median *Dc* (1.8118) being statistically indistinguishable from its clustered null median (1.8257; *p* = 0.285) (Table S3), indicating that IC’s nuclear spatial distribution could probably be fully accounted for by a two-level stochastic clustering process. Nonetheless, FEA showed the most extreme departure from CSR, with the largest gap between real (1.6881) and CSR (1.8425) *Dc* values of all subtypes (*p* = 3.96 × 10^−115^), while simultaneously remaining significantly below the clustered null (*p* = 8.03 × 10^−23^). Taken together, the results identified FEA as the subtype furthest from both stochastic reference processes simultaneously, and IC as the subtype whose spatial organisation is indistinguishable from elementary stochastic clustering, establishing a non-monotonic trajectory of organisational surplus across the pathological spectrum that does not follow the linear progression hypothesis.

### 3.3. Metric Complementarity

The investigation into metric complementarity confirmed that *Dm* and *Dc* capture distinct spatial features of the tissue landscape. Pearson correlation analysis showed a moderate linear relationship (*r*^2^ = 0.31, *p* < 0.001) (Fig. 2(b-c)), indicating that the morphological complexity of nuclear boundaries and nuclear spatial correlation provide 69% independent geometric information regarding the architecture, justifying their combined use in a multivariable diagnostic framework. The observation was supported by the Bland-Altman analysis, which demonstrated minimal systematic bias between the descriptors (*mean difference* = 0.015, *limit of agreement* = ±0.12), while the concordance correlation coefficient (*CCC* = 0.456) and the variance inflation factor (*VIF* = 1.45) further validated the absence of multicollinearity (Fig. S1.1).

### 3.4. Inter-subtype discrimination and pathological transitions

Both the FDs exhibited significant sensitivity to the breakdown of relational order or null progression across the pathological subtypes from the Kruskal-Wallis test (*p* < 0.001) with a medium to medium-large effect size for *Dm* and *Dc*, respectively. The Kruskal-Wallis test was employed due to violations of normality, assessed by the Shapiro-Wilk test (*p* < 0.001) for all subtypes and significant homogeneity of variance, assessed by Levene’s test with *Dc* (*p* = 5.5 × 10^−13^) and *Dm* (*p* = 1.2 × 10^−53^), respectively.

The mean rank concerning *Dc* follows FEA < N < PB < UDH ≈ ADH < IC < DCIS, identifying FEA as the subtype with the most spatially condensed nuclear distribution and DCIS with the most dispersed (Table S1). For *Dm*, the mean rank follows N < IC < PB < DCIS < ADH < UDH < FEA, identifying normal tissue as having the lowest boundary complexity and FEA the highest (Fig. 3(a), S1.2). The dissociation between the two rank orderings for FEA, i.e., the lowest *Dc* but highest *Dm* reflects its unique position in the configuration space of breast tissue architecture in carcinoma progression. Grounded in the null model results, FEA occupies the most extreme position relative to both stochastic reference processes, simultaneously exhibiting the most condensed nuclear spatial distribution and the most complex boundary morphology of any subtype.

**Fig. 3.**
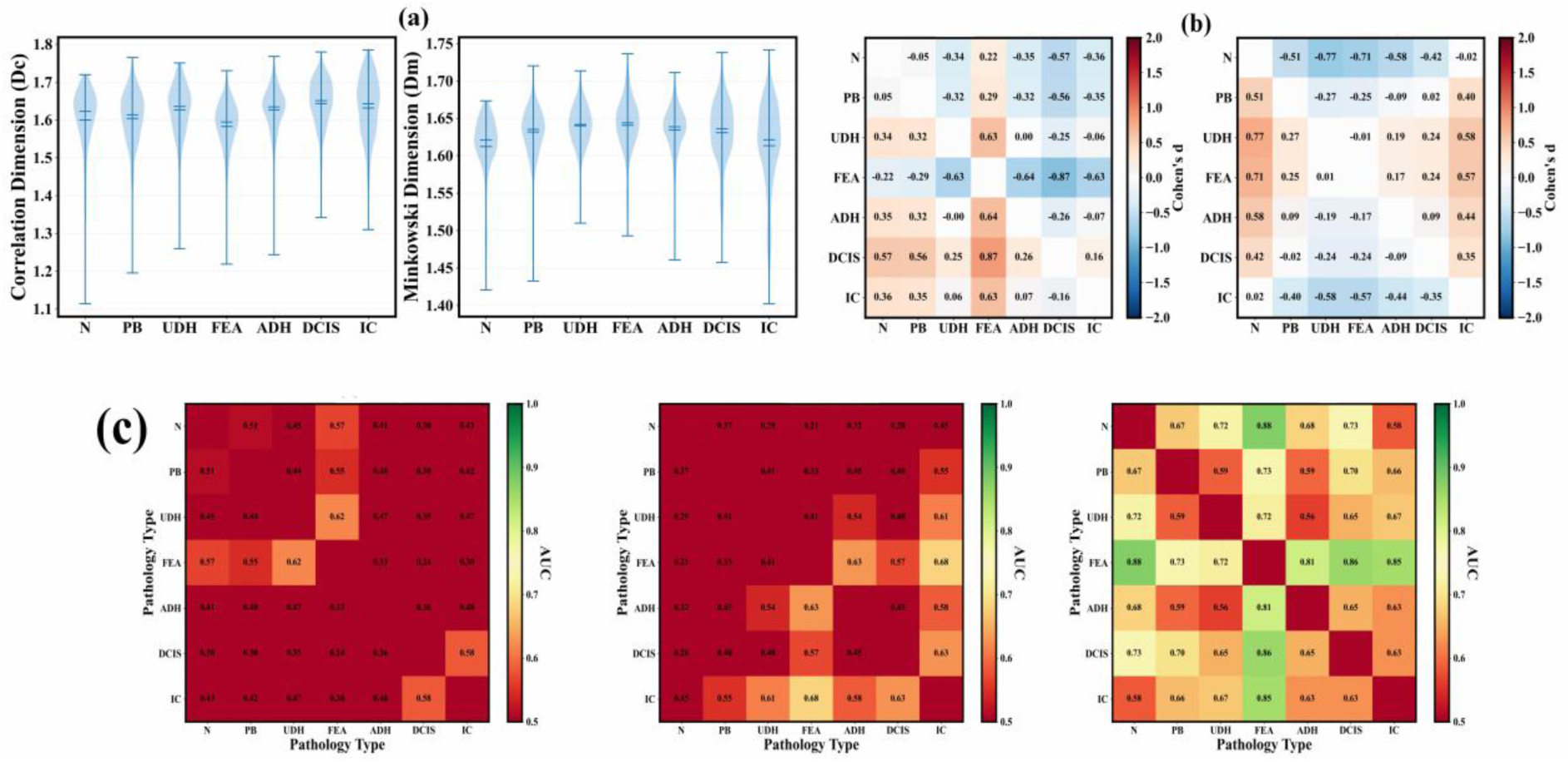
**(a)** Distribution of Correlation Dimension (Dc) and Minkowski Dimension (Dm) across subtypes; FEA shows the lowest and highest value, respectively, **(b)** Pairwise Cohen’s d heatmaps for Dc (left) and Dm (right) across 21 comparisons. Color scale ranges from blue (large negative effect, d ≈ −2) through white (no effect, d ≈ 0) to red (large positive effect, d ≈ 2), and **(c)** AUC matrices for transitions, evaluated at the WSI level (n = 368), using three feature sets: Dc (left), Dm (middle), and Dc + Dm combined (right). Color scale indicates discrimination performance from dark red (AUC ≈ 0.5, no discrimination) through yellow/orange (moderate, AUC ≈ 0.6-0.7) to green (good discrimination, AUC ≈ 0.8-0.9). Combined features achieve the highest discrimination (mean AUC = 0.677) compared to individual dimensions (Dc: 0.474, Dm: 0.483). Best transitions include N→FEA (0.878) and FEA→ADH (0.815)

Furthermore, pairwise discrimination analysis of 21 pathological transitions revealed distinct performance profiles for individual and combined fractal features (Fig. 3(b)). While individual dimensions achieved modest mean area under the curve (AUC) values (*Dc* = 0.474, *Dm* = 0.483), with *Dm* demonstrated superior statistical robustness, 8/21 transitions remaining significant after Bonferroni correction (α = 0.00238) compared to 4/21 for *Dc*, increasing to 12/21 and 10/21, respectively, under false discovery rate (FDR) correction. The combined feature set (*Dc* + *Dm*) substantially enhanced discriminative capacity, achieving a mean AUC of 0.677 (Fig. 3(c)), with peak performance, transition from the N to the FEA subtype (*AUC* = 0.878) and FEA to ADH subtype (*AUC* = 0.815) (Fig. 4(a-c)). However, this came at the cost of statistical robustness, i.e., no transitions reached significance after multiple testing correction (0/21 Bonferroni, 0/21 FDR), reflecting the reduced statistical power inherent to multivariate models with limited sample sizes. In addition, for clinically critical transitions, performance was moderate, ADH→DCIS (*AUC* = 0.649) and DCIS→IC (*AUC* = 0.626). Notably, the modest DCIS→IC discriminability is consistent with the null model finding that IC’s correlation dimension is statistically indistinguishable from a clustered null, highlighting that the transition from DCIS to IC subtype represents not merely a quantitative shift in fractal parameters but a qualitative change in the governing spatial process, which WSI-level averaging may partially obscure. For this analysis, we focused on the 320 WSIs containing multiple ROIs, excluding 48 single-ROI samples. These 320 WSIs comprised 84 pure-pathology samples (all ROIs with identical subtypes) and 236 mixed-pathology samples (ROIs with different subtypes).

**Fig. 4.**
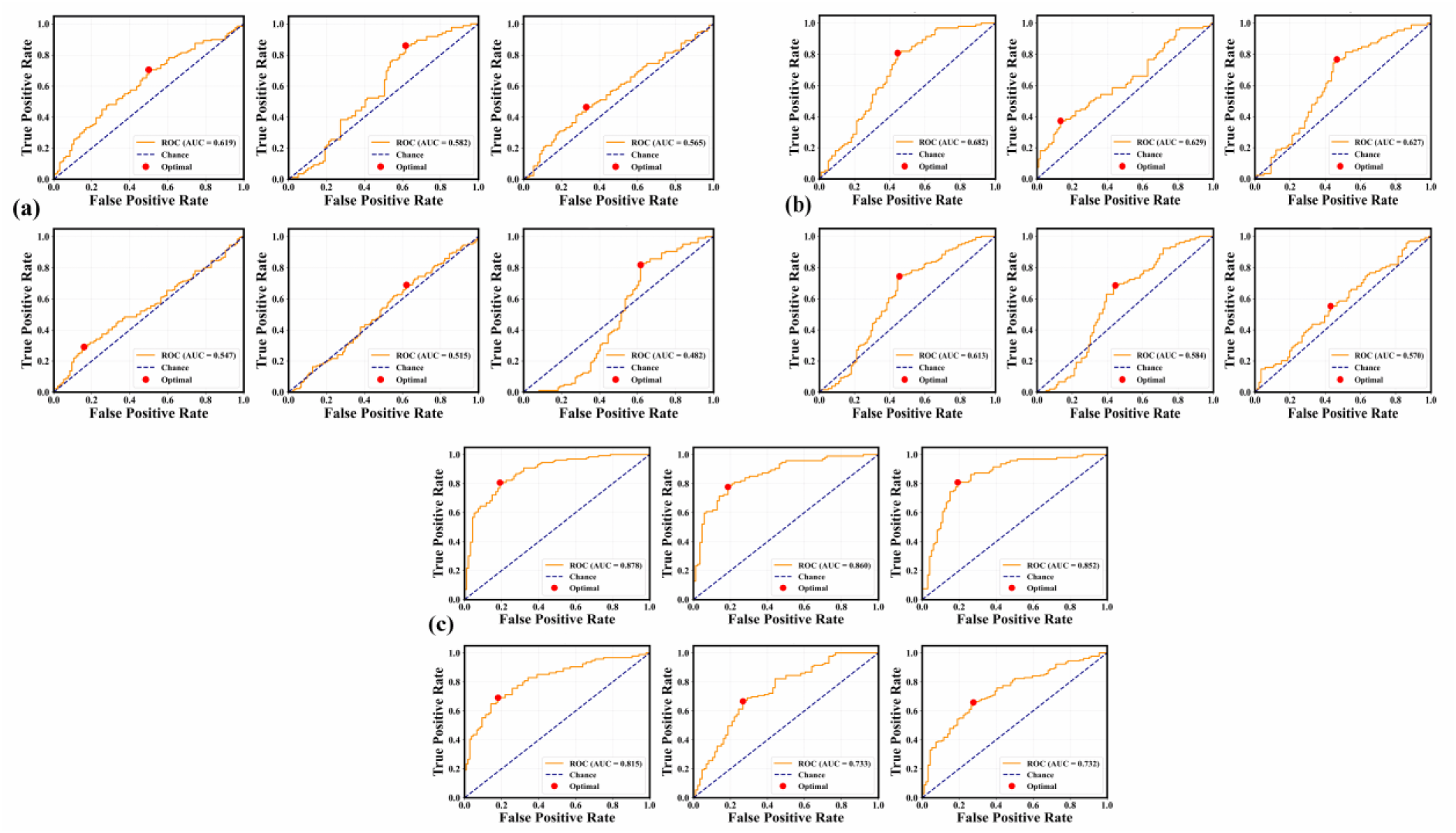
ROC analysis for representative pairwise transitions across the null pathological progression evaluated at the WSI level. **(a)** General (non-adjacent) transitions showing varying discrimination (N→FEA: AUC=0.878; UDH→FEA: 0.619; FEA→IC: 0.682), **(b)** Adjacent transitions with moderate performance (FEA→ADH: 0.815; ADH→DCIS: 0.649), and **(c)** Critical clinical transitions including DCIS→IC (AUC=0.626). Adjacent transitions show lower discrimination compared to general transitions due to smaller architectural differences between consecutive pathological stages. Blue dashed diagonal line represents chance performance (AUC=0.5). Red circle indicates the optimal threshold determined by Youden’s index *J* = *Sensitivity* + *Specificity* − 1.

### 3.5. Intra-tumor spatial heterogeneity

To account for the stochastic nature of carcinogenesis, intraclass correlation coefficients (*ICC*) were used to partition spatial variance within the WSI-level or to check the degree of heterogeneity within the 320 WSIs. The *ICC* for *Dc* was 0.469, indicating that 53.1% of the observed variance in global spatial organization occurs within the same WSI, highlighting significant intratumoral heterogeneity. In contrast, *Dm* exhibited a higher *ICC* of 0.713 (Fig. 5(a)), suggesting that local morphological boundary complexity is more consistent within a given sample than global spatial organization. Further analysis of spatial variance partitioning demonstrated that mixed-pathology WSIs exhibit 52% higher heterogeneity than pure-pathology samples, characterized by mean coefficients of variation (*CV*) of 2.92 and 1.92, respectively. The observed disparity in architecture was confirmed as statistically significant through an independent two-sample *t*-test on the within-WSI standard deviation (*t* = 5.255, *p* < 0.001) (Fig. 5(b)), demonstrating that spatial heterogeneity reflects the co-existence of multiple subtype states rather than intrinsic pathology characteristics. One-way ANOVA revealed significant differences in absolute within-WSI variance across subtypes for both *Dc* (*F* = 12.65, *p* < 0.001, *η*^2^ = 0.508) and *Dm* (*F* = 33.89, *p* < 0.001, *η*^2^ = 0.734). However, when heterogeneity was normalized using the *CV*, differences became non-significant (*F* = 1.547, *p* = 0.162, *η*^2^ = 0.029) (Fig. 5(c)), indicating that while absolute variance differs, relative heterogeneity remains proportional to mean FD values across subtypes. Additionally, the within-WSI or intratumoral heterogeneity was observed to significantly and positively correlate with ROI sampling density, as shown in Fig. 5(d).

**Fig. 5.**
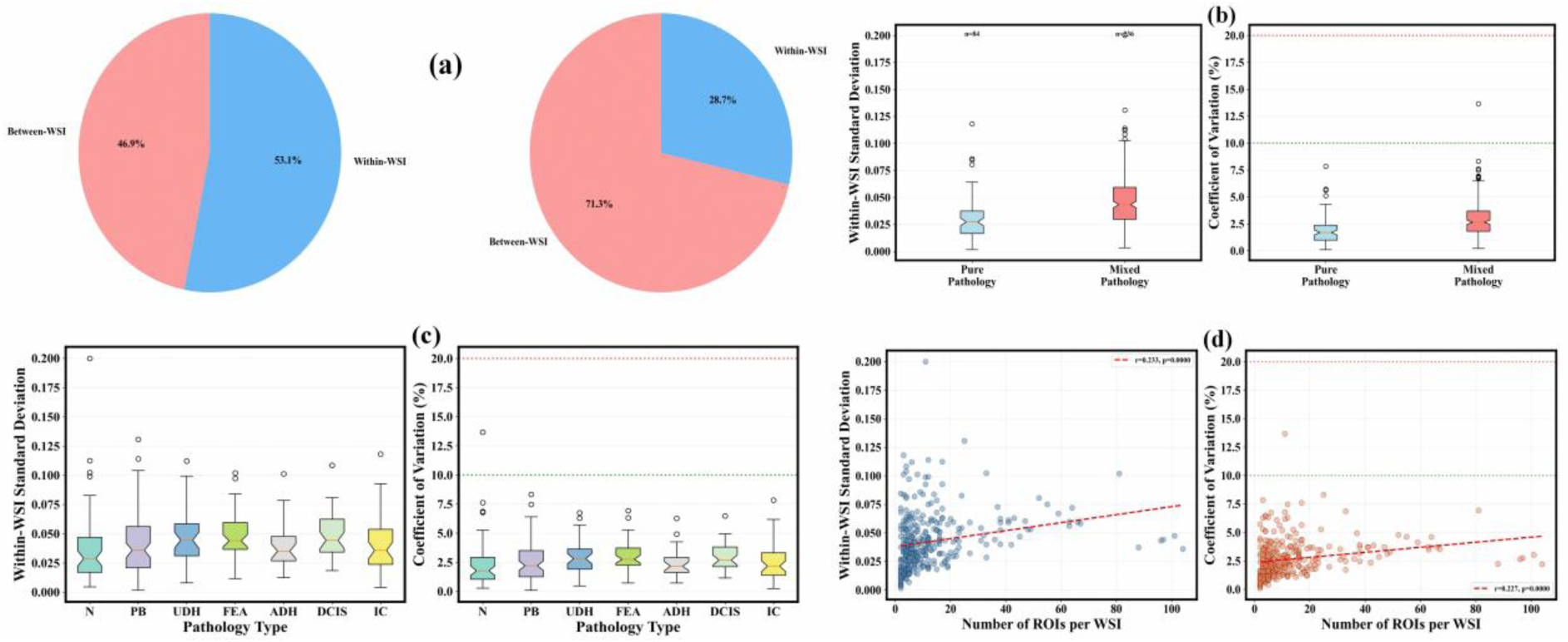
Variance decomposition and heterogeneity analysis across 368 WSIs, excluding 48 single-ROI WSIs. **(a)** ICC shows substantial within-WSI variance: Dc = 0.469 (53.1% within), Dm = 0.713 (28.7% within), **(b)** Mixed WSIs demonstrate 52% higher heterogeneity than pure; CV:2.92 vs. 1.92; independent *t*-test (t=5.255, p < 0.001). Red dashed lines indicate mean CV values, **(c)** Variance ANOVA: Dc F = 12.65, Dm F = 33.89 (both p < 0.001); CV non-significant (F = 1.547, p = 0.162), and **(d)** Heterogeneity correlates with ROI density (ρ = 0.400, p < 0.001). Red dashed lines indicate linear regression fits.

### 3.6. Confounding Analysis

Confounding analysis revealed distinct patterns for the two FDs. *Dc* demonstrated a strong correlation with scaling fit quality (*r* = 0.751, *ρ* = 0.892) and a significant correlation with measurement uncertainty (*r* = −0.847, *ρ* = −0.86) (Fig. 6(a-b)). This confounding remained consistent across pathologies (*r* = 0.763 − 0.834), indicating a systematic methodological relationship between the dimension estimate and the quality of the power-law behavior rather than biological variation specific to any subtype. Conversely, *Dm* showed weak correlation with fit quality (*r* = −0.099, *ρ* = 0.091, *p* < 0.001) (Fig. 6(c)) and moderate correlation with uncertainty (*r* = −0.541, *ρ* = −0.644, *p* < 0.001). The *Dm* − *R*^2^ relationship varied by pathology with *r* ranging from −0.431 in the N subtype to 0.357 in the UDH subtype, suggesting that any residual confounding in *Dm* is pathology-dependent rather than systematic. Multivariate regression controlling for *R*^2^, standard error, and pathology explained 73.9% of the variance in *Dc* and 94.4% in *Dm*. For *Dc*, both *R*^2^ (*t* = 116.3) and error (*t* = −89.7) were dominant predictors (*p* < 0.001) while for *Dm*, fit quality, and uncertainty again dominated (*t* > 400, *p* < 0.001), with pathology contributing modest but significant effects. The significantly strong negative correlation between *R*^2^and standard error (*r* = −0.916) confirmed that better fits yield more precise dimension estimates (Fig. 6(d)). The observed patterns are consistent with the physical interpretation that *Dc*, being estimated from a point-correlation integral across a wide spatial range, is more sensitive to global scaling regularity and therefore to the quality of the power-law approximation, while *Dm*, estimated over a narrow sub-decade scaling range, is governed more by local boundary geometry than by the global regularity of the fit.

**Fig. 6.**
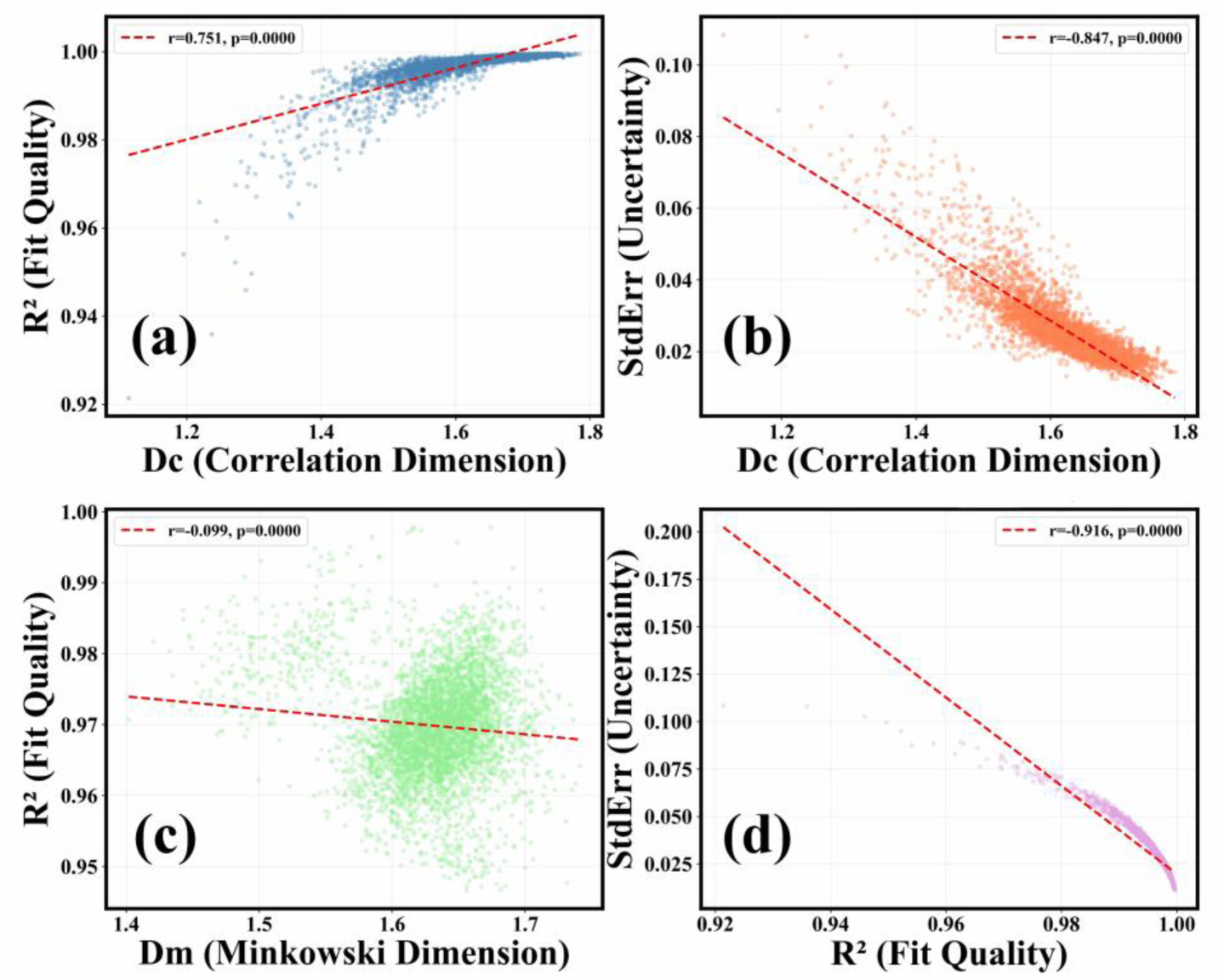
(a) Dc vs fit quality (R²): strong positive confounding (r = 0.751, p < 0.001). (b) Dc vs standard error (StdErr): strong negative correlation (r = −0.847, p < 0.001). (c) Dm vs R²: minimal confounding (r = −0.099, p < 0.001), and (d) R^2^ vs StdErr: moderate correlation (r = - 0.916, p < 0.001). The red dashed line represents linear regression.

### 3.7. Classification Performance

The clinical utility of the multiscale fractal framework was evaluated through the comparative benchmarking of 12 distinct combinations of machine learning architectures and feature sets. We evaluated four classifiers, Logistic Regression, Random Forest, K-Nearest Neighbors (KNN), and Support Vector Machines (SVM), across three feature configurations, i.e., *Dc* only, *Dm* only, and the combined multiscale set. Stratified 5-fold cross-validation at the WSI level (*n* = 368) established a clear hierarchy of performance and ensured balanced class representation across folds. Also, in consideration of the moderate *ICC* for *Dc* and high *ICC* for *Dm*, WSI-level splitting prevented data leakage from multiple ROIs within the same patient sample. Subsequently, the SVM model emerged as the most robust classifier with an average *AUC* = 0.62, followed by random forest (= 0.612) and KNN (= 0.6), while logistic regression (= 0.567) demonstrated the least capacity to model the non-linear architectural transitions. Feature analysis substantiated the hypothesis of complementarity, with the multiscale combined feature set yielding the highest predictive power, with an average *AUC* = 0.635, outperforming *Dc* only (= 0.627) and significantly exceeding *Dm* only (= 0.538).

The top-performing combination for binary classification (Benign + Atypical vs. Malignant) was the random forest model utilizing both FDs, achieving an *AUC* of 0.673 ± 0.018, accuracy of 0.728 ± 0.019, and the F1-score of 0.329 ± 0.073. For ternary classification (Benign/Atypical/Malignant), the SVM model with *Dc* only provided the highest accuracy and F1-score of 0.549 ± 0.036 and 0.434 ± 0.043 (Fig. 7(a-b)), respectively, suggesting that the fractal descriptor measuring both the local nuclear centroid correlation or clustering and global connectivity can serve as a primary driver in distinguishing broad pathological categories. The observed performance indicates that FDs of nuclear spatial architecture carry a statistically significant architectural signal, motivating their integration into computational pathology pipelines, but are insufficient as standalone clinical diagnostic tools.

**Fig. 7.**
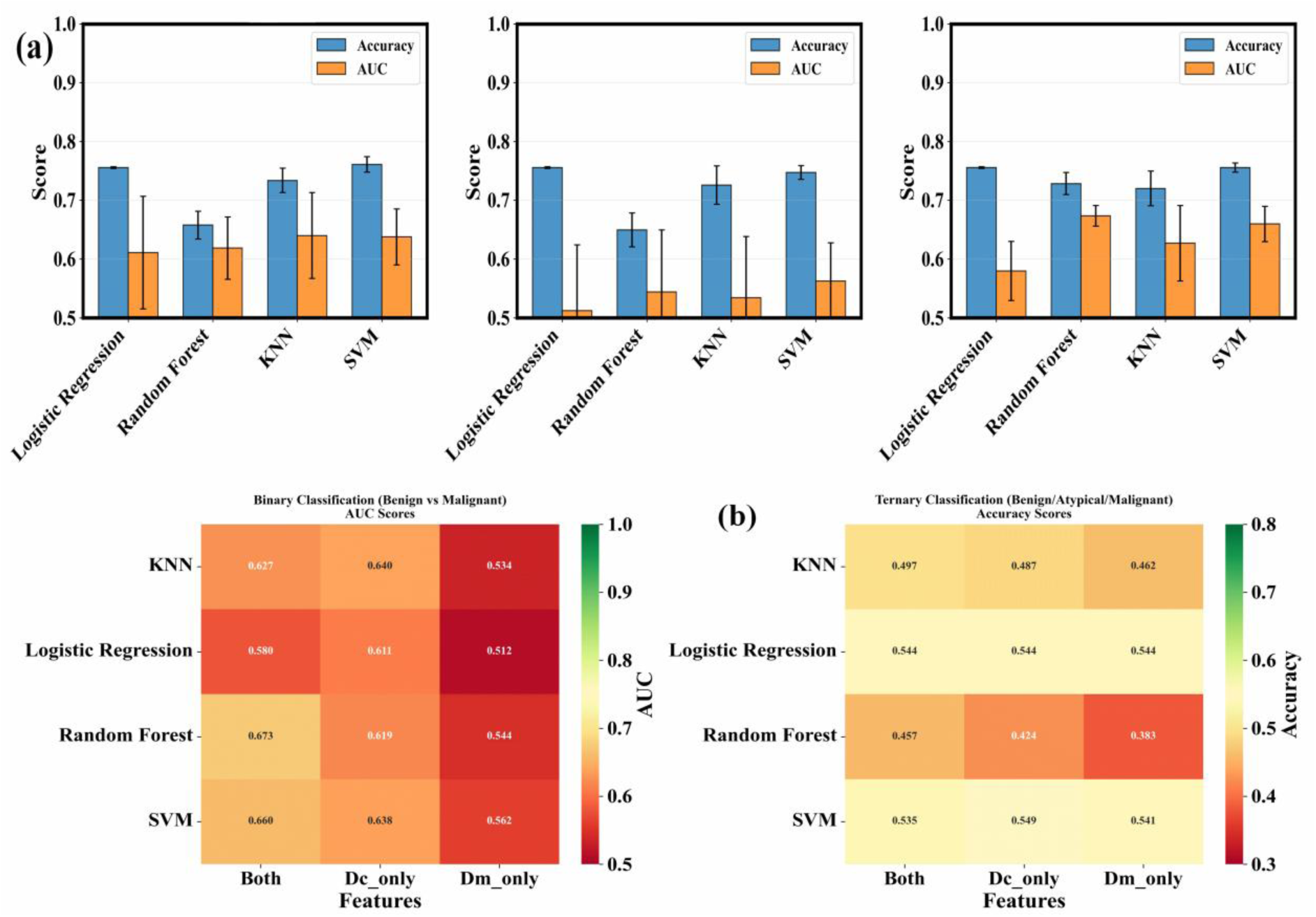
**(a)** Classifier performance across feature sets: Random Forest + Both achieves best AUC = 0.673±0.018 (binary); SVM + Dc best for ternary (0.549), indicated by error bars representing standard deviation across the 5 folds, and **(b)** Performance heatmaps showing combined features consistently outperform individual dimensions

### 3.8. Spatially Extreme Fractal Signatures

To account for the localized worst region effects characteristic of breast carcinogenesis, we extracted a comprehensive fractal signature per WSI, including the Maximum, Minimum, Range, and Mean of both *Dc* and *Dm*. This analysis aimed to determine if localized aggressive attractors (extreme values) serve as superior predictors of malignancy compared to global heterogeneity or WSI-level averages. Comparative ROC analysis identified *Dc*_*max* as the best-performing single predictor of malignancy (*AUC* = 0.59) though predictive power remained modest and only marginally better than the WSI-level average, narrowly outperforming the WSI-level average (*Dc*_*mean*, *AUC* = 0.588) and the combined maximum signature (*AUC* = 0.564) (Fig. 8(a)). Heterogeneity metrics, such as *Dm*_*range* (= 0.546) and *Dc*_*range* (= 0.509), demonstrated lower predictive utility.

**Fig. 8.**
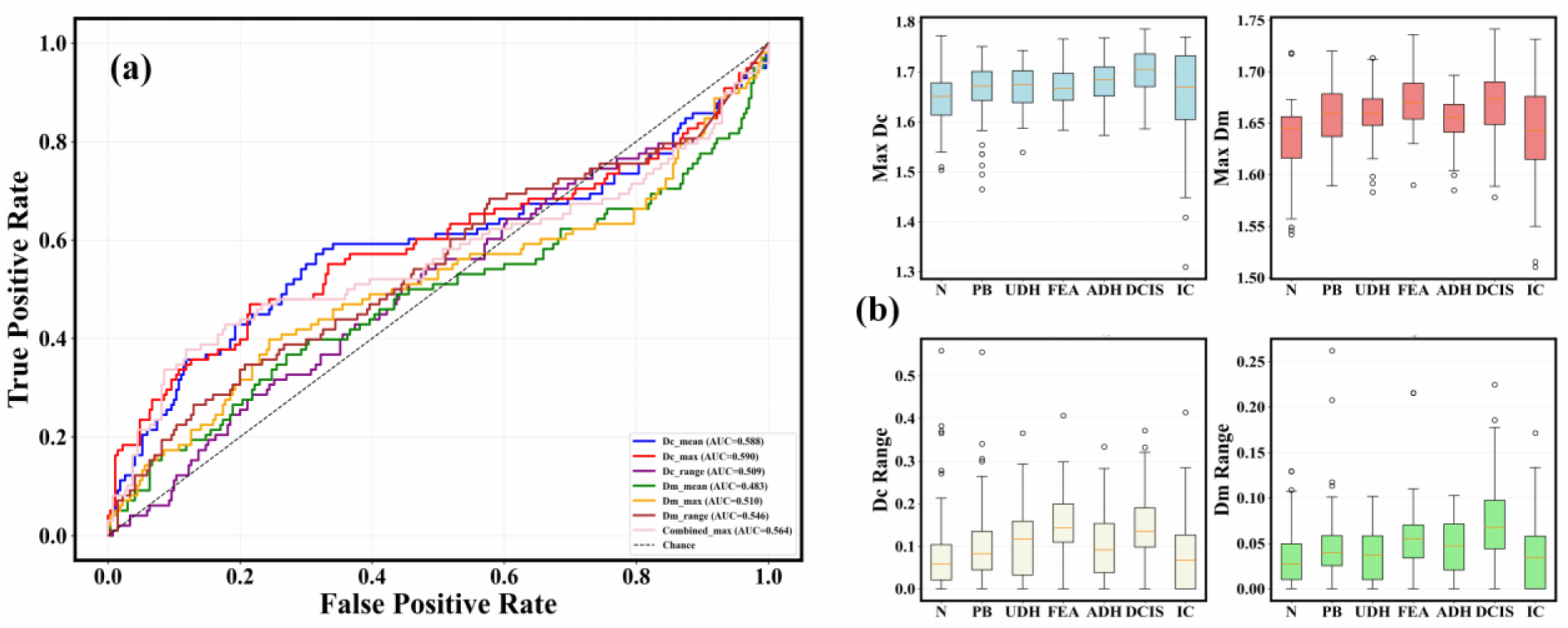
Analysis of localized fractal signatures (maximum, minimum, range, and mean values per WSI) to test whether extreme architectural features predict malignancy better than WSI-level averages. **(a)** ROC curves for eight signature metrics showing limited discrimination (best: Dc_max AUC=0.59), and **(b)** Maximum and range distributions by pathology; Dc_max F=4.34 (p=0.0003) (top-left panel, blue box), Dm_max F=8.31 (p<0.0001) (top-right panel, red box). Bottom panels display Dc_range (bottom-left, yellow box) and Dm_range (bottom-right, green box) showing within-WSI heterogeneity magnitude. Mixed WSIs show higher extremes (Dc_max: t=6.76, Dm_max: t=5.45, both p<0.0001). Red dashed horizontal lines indicate mean values, and outliers (circles) represent WSIs with extreme architectural features.

Furthermore, we observed a significant spatial dependency, i.e., mixed-pathology WSIs exhibited significantly higher maximum FDs than pure-pathology ones. For *Dc*, mixed-WSI *Dc*_*max* (= 1.682) was significantly higher than pure-WSI (= 1.639) with *t* = 6.76, *p* < 0.0001. A similar trend was also observed for *Dm* (1.661 *vs*. 1.64) with *t* = 5.45, *p* < 0.0001 (Fig. 8(b)). While these results confirm that spatial heterogeneity, represented by mixed lesion types, drives higher localized architectural disorder, the overall predictive power for malignancy remained limited (*AUC* = 0.59), indicating that fractal signatures of spatially extreme regions are distributed across the tissue field rather than concentrated in focal high-disorder regions, consistent with the systems-level reorganisation of tissue spatial processes.

## 4. Discussion

This study investigates whether fractal-like nuclear spatial organisation in breast tissue constitutes a physically interpretable signature of pathological progression. By comparing FD estimates against null spatial models across seven subtypes, we show that the deviation of nuclear spatial patterns from stochastic baselines varies non-monotonically with pathological stage, with FEA exhibiting a unique directional dissociation between the two dimensions and IC showing dual convergence toward stochastic baselines. These observations are interpreted within a percolation-theoretic framework. Throughout, it is emphasised that *Dc* and *Dm* are measurements of nuclear spatial patterns, interpreted from centroid distribution and boundary morphology, respectively, and that connections to tissue-level biology are interpretive inferences consistent with the data rather than direct measurements.

### 4.1. Null model deviation as the operative biophysical quantity

An FD value in the range 1 < *FD* < 2 is a necessary but not sufficient condition for fractal-like behaviour, since any geometrically irregular boundary or non-random point pattern satisfies this criterion. In this regard, the physically meaningful quantity is therefore the deviation of the observed dimension from a stochastic reference process with known, non-fractal properties. All seven subtypes maintain nuclear spatial organisation significantly above the CSR baseline, confirming that no subtype exhibits spatially random nuclear arrangement. However, the comparison with the clustered null reveals a critical asymmetry, i.e., six subtypes show *Dc* significantly below the clustered null, indicating nuclear spatial organisation beyond what elementary aggregation accounts for, while IC alone does not (*p* = 0.285). For *Dm*, the Cohen’s *d* of the paired real-versus-null difference declines from PB (1.475) through DCIS (0.832) to IC (0.423), tracking the progressive weakening of nuclear boundary complexity excess above the smooth ellipse baseline. Together, these results identify a non-monotonic trajectory of organisational surplus that does not follow the histopathology-defined linear clinical progression sequence and motivates a regime-based rather than gradient-based interpretation.

### 4.2. Physical boundary conditions and subtype-specific fractal-like signatures

The histological descriptions of each subtype in the BRACS dataset [3] provide a basis for interpreting the observed nuclear spatial patterns, with the caveat that the reported fractal descriptors measure nuclear geometry and distribution, not the cellular or tissue-level processes themselves. The following interpretations are therefore offered as geometric analogues that are consistent with the known histology, rather than as direct evidence of those histological processes.

Normal tissue, characterised by a dual-layer epithelial architecture [28], shows substantial *Dm* null deviation, geometrically consistent with a nuclear population containing morphologically diverse boundary shapes. Its *Dc* lies between the two null models, consistent with a nuclear distribution that is neither random nor maximally condensed, a pattern compatible with spatially distributed nuclei across a structured architectural field. The UDH subtype, described histologically as a cohesive but disorderly proliferation of oriented cells [3], shows intermediate values on both dimensions and moderate null deviations, consistent with an intermediate architectural state in which neither maximal boundary complexity nor maximal spatial condensation is reached. The intermediate position of UDH in both analyses is consistent with a nuclear arrangement that partially departs from the normal pattern without reaching the extremes characteristic of atypical subtypes.

PB shows the largest *Dm* null deviation of all subtypes (*d* = 1.475) and *Dc* below the clustered null. PB is a heterogeneous diagnostic category encompassing multiple architecturally distinct lesion types [3], and its extreme *Dm* excess most likely reflects the wide range of nuclear boundary morphologies present across this diversity of lesion types rather than a single coherent architectural state. This compositional heterogeneity should be considered when interpreting PB’s fractal position, and its classification performance should be interpreted with corresponding caution.

### 4.3. The FEA dissociation and the decoupling of spatial scales

FEA consistently occupies the lowest *Dc* position across both the ComsystanJ and null model analyses, with its nuclear centroid distribution is more condensed than any other subtype and departs furthest from the CSR null. Simultaneously, FEA shows high *Dm* null deviation (*d* = 1.096), placing it in the upper range of boundary complexity surplus across both analyses, though ranking varies by implementation. The ComsystanJ-based analysis places FEA highest in *Dm* mean rank, while the null model real medians place UDH highest and normal second, with FEA third. Nonetheless, a consistent observation across both analyses is that FEA combines the lowest *Dc* with high *Dm*, a directional dissociation not observed in any other subtype.

The dissociation indicates that the FEA subtype is characterised simultaneously by nuclear centroids that are more spatially condensed than any other subtype and by nuclear boundaries that are more morphologically complex than advanced atypical and invasive subtypes. These two geometric observations are in opposite directional senses relative to the rest of the spectrum. FEA’s histological definition, including loss of orientation, flat proliferative growth, and low-grade cytological atypia [3,29–30], is geometrically consistent with a nuclear spatial pattern in which positional freedom is reduced (consistent with the low *Dc*) while individual nuclear boundary irregularity is increased (consistent with the high *Dm*). However, it should be noted that the fractal measurements do not directly correlate with cell polarity, basement membrane organisation, or cytological atypia; the connection between these histological features and the observed nuclear geometry is an inference. A possible way of investigating this connection is by studying directly co-registered histological and fractal measurements. The geometric dissociation itself, i.e., the co-occurrence of low *Dc* and high *Dm*, is the empirical finding; its biological interpretation is a hypothesis consistent with the known histology.

### 4.4. Invasive carcinoma and progressive weakening of nuclear spatial organization

IC shows the weakest *Dm* null deviation of all subtypes (*d* = 0.423), less than half that of DCIS at (*d* = 0.832) and is the only subtype whose *Dc* is statistically indistinguishable from the clustered null (*p* = 0.285). Both FDs thus converge toward their stochastic baselines simultaneously at IC, a dual convergence not observed at any other stage. The histopathological definition of IC centres on loss of peripheral myoepithelial cells and stromal invasion [31–32], and the observed nuclear spatial patterns are geometrically consistent with a nuclear arrangement that, at both the centroid distribution scale and the boundary morphology scale, no longer departs meaningfully from stochastic processes. However, whether this convergence is mechanistically linked to myoepithelial loss, changes in nuclear mechanical properties [33,34], or other aspects of the invasive transition cannot be determined from the nuclear geometry alone; the fractal observations are compatible with these mechanisms, but their correlation or mechanistic link has not been reported. Nevertheless, nuclear spatial organisation as measured by *Dc* and *Dm* is weaker at IC than at any other stage, and that this weakness is captured by the proximity to stochastic null baselines rather than by the absolute FD values alone.

### 4.5. A percolation-theoretic framework for the non-monotonic trajectory

The non-monotonic trajectory of null model deviations across the seven subtypes, i.e., high at early stages, maximal at atypical transitional stages, then declining through in-situ to invasive carcinoma, is qualitatively consistent with the predictions of percolation theory applied to a system undergoing structural regime transitions [35–36]. In percolation theory, fractal-like, scale-invariant behaviour emerges near critical transitions when no dominant characteristic length scale governs the system, while subcritical regimes (characterised by a single dominant structural scale) and supercritical regimes (where a spanning structure re-establishes a characteristic scale) show weaker fractal signatures [37–38]. This provides a physical framework for interpreting the observed AUC profile and null deviation trajectory. It is noted that the following regime mapping is drawn from the combined evidence of both the ComsystanJ and null model analyses. Where the two analyses are consistent, the regime assignment is stated directly; where they differ, the discrepancy is noted explicitly, and the assignment is qualified accordingly.

Normal tissue shows high *Dm* null deviation and an intermediate-to-low *Dc* position, i.e., second lowest in the ComsystanJ mean rank ordering, but fourth of seven in the null model real median ordering (1.7985). The two analyses, therefore, partially disagree on the normal tissue’s *Dc* rank, likely reflecting the algorithmic and parametric differences between implementations. Notably, this disagreement is not an artefact of distributional asymmetry since replacing the subtype mean with the median ranks moves PB below N in *Dc*, aligning with the null model ordering, while N and UDH retain their reversed positions across the two implementations, confirming that the residual discrepancy reflects implementation differences rather than sensitivity to descriptive statistic choice (Table S1). On average, N subtype nuclear spatial patterns show a substantial departure from both null baselines on *Dm* and moderate-to-high departure from CSR on *Dc*, consistent with a well-structured nuclear arrangement compatible with a subcritical regime interpretation. However, it is acknowledged that its position in the *Dc* space is less definitively low in the null model analysis.

PB presents an ambiguity that neither analysis resolves consistently. Its *Dm* null deviation is the largest of all subtypes (*d* = 1.475), which by itself would place it firmly in a high-surplus, subcritical regime. However, the median *Dc* for the real images in the null model analysis is the second lowest of all subtypes (1.7424), falling significantly below the clustered null (*p* = 5.2 × 10^−8^), a position that geometrically overlaps with the critical transitional regime rather than the subcritical one. ComsystanJ places PB third lowest in *Dc* mean rank, which is directionally consistent but less extreme. This inconsistency is not resolvable from the investigated data and most likely reflects the compositional heterogeneity of the PB subtype, which encompasses multiple architecturally distinct lesion types [3] rather than a single coherent nuclear spatial state. PB’s extreme *Dm* excess, combined with its low *Dc* position, may therefore represent a superposition of distinct nuclear spatial patterns from its constituent lesion types rather than a physically unified regime. Subsequently, its placement in the percolation mapping should be treated as ambiguous, overlapping both subcritical and transitional regime characteristics, rather than definitively assigned to either.

UDH shows moderate null deviations on both FDs and an intermediate position in both ranking analyses, consistent with an early transitional state in which nuclear spatial patterns partially depart from the structured normal arrangement without reaching the extremes characteristic of the atypical subtypes [3]. Both analyses agree on this intermediate placement, making it the most consistently assigned regime in the spectrum.

FEA and ADH occupy the critical transitional regime, though underpinned by a different rationale. FEA shows the lowest *Dc* across both analyses, indicating that its nuclear centroid distribution departs furthest from CSR and remains significantly below the clustered null, combined with high *Dm* null deviation. This directional dissociation between the two dimensions, unique to FEA, is consistent with a nuclear spatial state in which no single scale dominates organisation simultaneously at the centroid distribution and boundary morphology levels, the condition under which fractal-like behaviour is predicted to be most pronounced in a percolation-theoretic framework. ADH’s transitional assignment rests primarily on the statistical significance of its departure from the clustered null in *Dc* and its moderate *Dm* excess, both consistent with a nuclear arrangement that departs beyond stochastic clustering. However, ADH’s absolute *Dc* rank in the null model analysis (fifth of seven, real median 1.8047) is near those of DCIS and IC, which are mapped to the supercritical regime. The transitional assignment for ADH is therefore supported by the magnitude of its null model departure rather than by its absolute *Dc* position.

DCIS represents an intermediate-to-supercritical transition, i.e., its *Dc* in the null model analysis is among the highest values and approaches the CSR null from below, while its *Dm* null deviation declines. Both analyses place DCIS near the upper end of *Dc* space, consistent with a nuclear centroid distribution that is approaching spatial uniformity and in which a characteristic distribution scale is beginning to re-emerge. IC represents the fully supercritical regime where both FDs converge toward their respective stochastic baselines, with *Dc* statistically indistinguishable from the clustered null and *Dm* Cohen’s d the lowest of all subtypes. This convergence is consistent across both analyses.

Notably, this framework reframes the modest DCIS→IC classification AUC (0.626) not as a methodological limitation but as a physically expected consequence of regime structure. Once the transition into the supercritical invasive regime is complete, the remaining fractal information is minimal, i.e., the system has already converged toward stochasticity, and there is relatively little organisational structure left to discriminate. Conversely, the high AUC for N→FEA and FEA→ADH reflects the large separation in nuclear configuration space between the subcritical and critical regimes, where null model deviations are most divergent. Nevertheless, in biological systems, transitions occur as gradual crossovers rather than sharp critical thresholds [39], and the finite scaling range of both FD estimates precludes any claim of exact critical exponents or universal scaling behaviour; the percolation framework describes only the qualitative regime structure. Also, the regime mapping relies solely on nuclear spatial measurements, and whether the inferred regime transitions correspond to distinct tissue-level structural transitions requires independent validation beyond the scope of this study.

### 4.6. Methodological Considerations

The strong confounding of *Dc* by its own scaling fit quality, consistent across all subtypes, indicates that *Dc* estimates partially encode the regularity of the power-law approximation rather than exclusively the underlying nuclear spatial geometry. *Dc* should therefore be interpreted with *R*^2^as a covariate, and future implementations should seek an expanded scaling range to reduce this coupling.

*Dm* shows negligible *R*^2^confounding and pathology-dependent rather than systematic confounding patterns, making it a more methodologically self-consistent signature. Nevertheless, a notable implementation-dependent discrepancy in *Dm* was observed for the N subtype. Specifically, ComsystanJ analysis places it lowest in *Dm* mean rank, while the null model implementation places it second highest in both raw median (1.3322) and effect size. This reversal most likely reflects the fixed versus adaptive scaling range difference between implementations. Normal tissue nuclei are characteristically smaller than those of atypical or malignant subtypes [40], and the adaptive *r*_*max*_in the null model implementation, set proportional to the median nuclear minor axis length per image, may more accurately confine estimation to the boundary-dominated scaling regime for small nuclei, avoiding the area-dominated regime at large dilation radii where *Dm* → 0. ComsystanJ’s fixed 10-step dilation sequence, applied uniformly regardless of nuclear size, may extend into this area-dominated regime for smaller nuclei, artificially depressing the relative *Dm* estimate for the N subtype. This ranking instability should be considered when interpreting subtype comparisons involving it, and it represents a significance for methodological reconciliation through matched scaling range experiments. This is reinforced by the comparison of subtype medians against mean ranks, which confirms that this discrepancy is not attributable to distributional skewness (Table S1).

Another basis of inter-implementation discrepancy, specific to *Dc*, arises from the algorithmic difference between the raster box scanning approach used in ComsystanJ and the Grassberger-Procaccia correlation integral used in the null model analysis. ComsystanJ partitions the image domain into grids of boxes at 11 sizes scaled by powers of two and approximates the correlation sum from squared pixel counts within each box, operating on pixelated binary images and inheriting sensitivity to image resolution and discrete grid structure. The null model implementation computes all pairwise Euclidean distances between nuclear centroid coordinates directly and evaluates the correlation integral over 15 geometrically spaced radii, with slope estimated by a sliding window maximising *R*^2^. These are related but non-identical estimators that converge theoretically but can diverge in practice at the limited scaling ranges available at standard histological resolution, particularly for subtypes with small nuclear populations or atypical spatial distributions [41,42]. The *Dc* ranking reversals between the two implementations, notably the Normal/PB exchange in the lower *Dc* range and the near-equivalence of DCIS and IC at the upper end, are likely attributable to this algorithmic difference as much as to any parametric difference, and should be considered alongside the fixed versus adaptive scaling range factor when assessing cross-implementation consistency. A matched algorithmic implementation across both primary analysis and null model comparison would provide a more rigorous basis for ranking stability assessment.

The observation from within-patient or intra-tumor heterogeneity analysis further indicates that single-ROI sampling is insufficient for stable *Dc* characterisation, with a minimum of three to five spatially distributed ROIs per WSI for each subtype for translational studies.

Finally, the fractal-like scaling shown by both descriptors can be characterised as statistical monofractal isotropic scaling within a bounded scaling range. *Dc* spans approximately one log-decade of scaling range, while *Dm* consistently spans a sub-decade range of approximately 0.78–1.0 log units, possibly inherent to histological nuclear resolution. It is noteworthy that both descriptors employ isotropic distance or dilation measures; the Euclidean pairwise distance for *Dc* and the disk-kernel dilation for *Dm*, highlighting that any directional anisotropy in nuclear arrangement or boundary morphology is averaged over rather than resolved. Self-affinity, which would imply distinct scaling exponents in different spatial directions, is therefore neither confirmed nor excluded by the present analysis, though it is biologically plausible for subtypes with oriented nuclear arrangement such as UDH. Similarly, the progressively declining *Dm R*^2^ with carcinoma progression is geometrically consistent with increasing scaling heterogeneity which would suggest the development of multifractal scaling in boundary morphological complexity in advanced subtypes. Nevertheless, the observed sub-decade scaling range is insufficient to distinguish true multifractal behaviour from approximate monofractal scaling.

### 4.7. Clinical implications and future directions

The combined fractal framework achieves AUC = 0.878 for the N→FEA transition and AUC = 0.815 for FEA→ADH, both involving lesions that are maximally undetectable by mammography and frequently underdiagnosed in core needle biopsies [43]. This can be interpreted from the unique position of FEA in the fractal configuration space as a maximally constrained, scale-decoupled system, suggesting that fractal analysis of nuclear spatial architecture is most valuable as a signature of the pre-malignant transitional state rather than as a malignancy classifier. The modest standalone classification performance (AUC = 0.673) positions these fractal dimensions as candidate auxiliary biomarkers within multifeatured diagnostic pipelines, where nuclear spatial architecture provides the scale-interaction component that individual nuclear morphometry cannot capture. Additionally, prospective validation in independent cohorts with multi-site ROI sampling is required for the validation regarding the significance of the reported FDs in clinical translation.

## 5. Conclusion

This study demonstrates that fractal-like nuclear spatial organisation across seven breast tissue subtypes, quantified through *Dc* and *Dm* validated against null spatial models, constitutes a physically grounded biophysical signature of pathological progression. The deviation above stochastic baselines, rather than absolute dimension values, is the operative quantity, establishing a non-monotonic organisational surplus trajectory. IC uniquely converges toward stochastic baselines on both dimensions simultaneously, while FEA exhibits a unique directional dissociation, the lowest *Dc* combined with high *Dm* null deviation, consistent with a geometrically interpretable decoupling of nuclear spatial organisation at the centroid distribution and boundary morphology scales. Interpreted within a percolation-theoretic framework, this trajectory maps onto qualitative subcritical, critical, and supercritical regime transitions, providing a physically grounded explanation for the observed peak and modest discrimination, respectively, as geometrically expected consequences of regime structure rather than methodological limitations. The scaling is characterised as statistical monofractal isotropic within a bounded range. *Dc* and *Dm* are positioned as candidate descriptors within multifeatured diagnostic pipelines, with particular utility for pre-malignant transitional states undetectable by standard breast imaging, pending prospective validation in independent cohorts with multi-site ROI sampling for clinical translation.

## Supplementary Information

The mathematical formulations and statistical rationale supporting the analyses, the Bland-Altman agreement analysis between *Dc* and *Dm* across all 4276 ROIs and stratified by pathological subtype, and the fractal dimensions trend across the pathological progression sequence are provided in the Supplementary Material (S1). The tables provide the full descriptive statistics of *Dc* and *Dm* estimated from ComsystanJ analysis across all seven subtypes. They also present the complete paired null model comparison results for *Dc* and *Dm*, respectively, including real and null medians, Wilcoxon signed-rank p-values, and Cohen’s d effect sizes per subtype.

## Data Availability Statement

The study used the publicly available BRACS dataset (https://www.bracs.icar.cnr.it/). The binary image data that support the findings of this study are available upon request from the corresponding authors. The custom-built programs and Fiji macros can be accessed from https://github.com/Abhi881991/Breast-Cancer-Histopathology-Nuclei-Images-Fractal-Analysis-and-Statistical-Analysis

## Acknowledgment

**A. D.** acknowledges the Department of Biotechnology (DBT)-India for the Research Associate fellowship vide Award Letter No. DBT-RA/2023/January/NE/3594.

**R. B.** acknowledges support from the Indo-French Centre for the Promotion of Advanced Research (69T08-2).

**M. K. J.** acknowledges support from the Param Hansa Philanthropies.

