## Supplementary Material for "Decoupling of spatial scales in breast pathology reveals fractal-like nuclear organization emergent from tissue spatial architecture"

#### Validation of Scaling Behavior:

1. The existence of fractal-like scaling was validated using the Coefficient of Determination ($R^2$):

$$R^2 = 1 - \frac{SS_{residual}}{SS_{total}}$$

Here, $SS$ means the sum of squares.

2. The Spearman test was applied to determine if the fit quality ($R^2$) declines with breast cancer progression.
3. The Shapiro-Wilk test was used to assess the distribution of the Minkowski dimension ($Dm$) and the Correlation dimension ($Dc$) values

#### Complementarity Analysis:

1. The linear relationship strength between $Dm$ and $Dc$ or their independence was measured using the Pearson correlation, where $r^2$ represents the shared variance.
2. Bland-Altman analysis was utilized to identify systematic bias and limits of agreement between the two dimensions.
3. Variance Inflation Factor (VIF) and Concordance Correlation Coefficient (CCC) were calculated to assess the need for the implemented multivariable approach and, subsequently, the degree of redundancy.

#### Pathology Discrimination

1. Due to the non-normal distribution of $Dm$ and $Dc$, the Kruskal-Wallis test was used to determine if the fractal measures differed across the subtypes.
2. Non-parametric effect size was calculated using $\varepsilon^2 = \frac{H-k+1}{N-k}$. Here, $k = 7$, $N = 4276$, and $H = \left[\frac{12}{N(N+1)} \sum \left(\frac{R_i^2}{n_i}\right)\right] - 3(N+1)$ is the Kruskal-Wallis test statistic.
3. Dunn's Post-hoc tests with Bonferroni correction were applied to identify specific pathological pairs with significant differences.
4. Cohen's d (pairwise): $d = \frac{(\mu_1 - \mu_2)}{\sqrt{[(n_1-1)\sigma_1^2 + (n_2-1)\sigma_2^2 / n_1 + n_2 - 2]}}$, was used to quantify the magnitude of effect between specific pairs.

#### Partitioning of Spatial Heterogeneity:

1. Intraclass Correlation Coefficients (ICC) were employed to partition variance between Whole Slide Images (WSIs) and Regions of Interest (ROIs): $ICC = \frac{\sigma_{between}^2}{(\sigma_{between}^2 + \sigma_{within}^2)}$
2. The Coefficient of Variation $CV = \left(\frac{\sigma}{\mu}\right) \times 100\%$ was calculated per WSI
3. Independent two-sample t-tests: $t = \frac{(\mu_1 - \mu_2)}{\sqrt{(\sigma_1^2/n_1 + \sigma_2^2/n_2)}}$ compared heterogeneity levels in mixed-pathology versus pure pathology cases

#### Machine Learning and Classification Performance:

1. Receiver Operating Characteristic (ROC) Analysis (Area Under the Curve (AUC)) was utilized to quantify the classification performance of 21 pairwise pathological transitions.
2. Youden's Index $(J)$ was used to find thresholds or optimal cut points that maximize sensitivity and specificity.
3. Hedge's $g$ was used for the effect size, ensuring robustness to small-sample and variance differences, in consideration of the lower sample size of WSIs.
4. Four classifiers (Logistic Regression, Random Forest, K-Nearest Neighbor (KNN), and Support Vector Machine (SVM)) were benchmarked using 5-fold cross-validation, utilizing AUC for binary classification and accuracy for ternary classification.
5. Compared the predictive power of different metrics, such as $Max\_Dc$, $Range\_Dm$, etc., to determine if localized extreme values or global heterogeneity better predict malignancy or worst region (DCIS or IC) than simple averaging.

### Figures

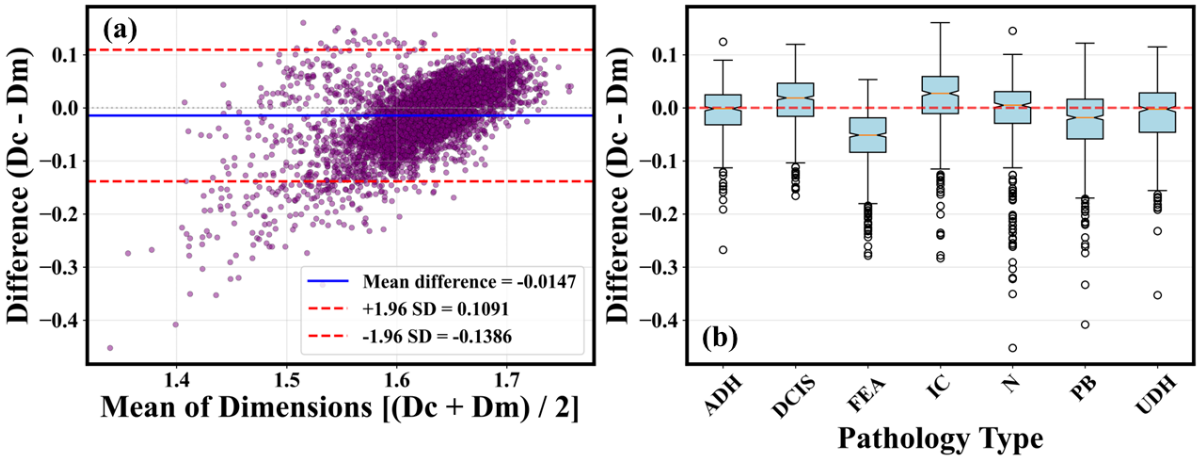


**Fig. S1.1. (a)** Bland-Altman plot for all ROIs (n = 4276) showing difference (Dc - Dm) versus mean [(Dc + Dm)/2]. Mean difference = -0.0147 (blue solid line) with limits of agreement ±1.96 SD = ±0.1091 (red dashed lines). Small systematic bias indicates Dm slightly exceeds Dc on average. Most points cluster near zero difference, confirming general agreement, and **(b)** Agreement by pathology type showing box plot distributions of differences for seven pathologies. FEA and IC show the largest negative differences (Dc < Dm), while ADH, DCIS, and N show near-zero median differences. The red dashed line at zero indicates perfect agreement. Variation in agreement across pathologies reflects biological differences in spatial architecture rather than measurement inconsistency.

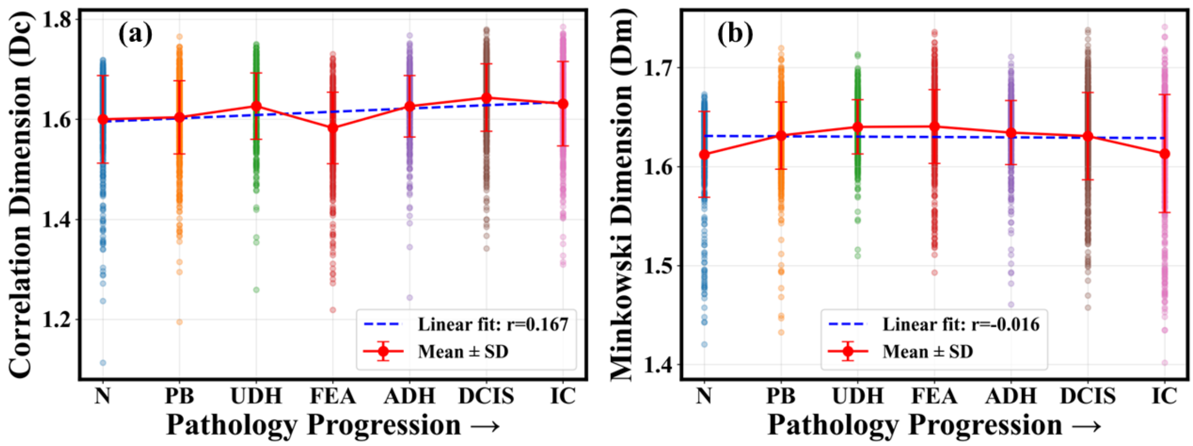


**Fig. S1.2. (a)** Dc across null progression showing non-monotonic trend. Linear fit (blue dashed, r = 0.167). The red line indicates mean ± standard deviation (SD) per pathology. FEA exhibits the lowest mean Dc (~1.59) while other pathologies cluster around 1.60-1.63, and **(b)** Dm across progression showing a weak declining trend (linear fit: r = -0.016). FEA shows the highest mean Dm (~1.64), inverting the pattern observed for Dc.

### Tables

**Table S1.** Descriptive statistics of the Correlation dimension (*Dc*) and Minkowski dimension (*Dm*) estimated from ComsystanJ analysis across seven breast tissue subtypes, ordered by clinical progression sequence. Values reported are mean, standard deviation, median, minimum, and maximum computed at the ROI level.

| Subtype (# ROIs) | ***Dc* (Mean)** | ***Dc* (Standard Deviation)** | ***Dc* (Median)** | ***Dm* (Mean)** | ***Dm* (Standard Deviation)** | ***Dm* (Median)** |
|---|---|---|---|---|---|---|
| N (476) | 1.6 | 0.0876 | 1.6230 | 1.6123 | 0.0434 | 1.6208 |
| PB (775) | 1.6040 | 0.0733 | 1.6135 | 1.6314 | 0.0339 | 1.6352 |
| UDH (507) | 1.6264 | 0.0665 | 1.6355 | 1.64 | 0.0274 | 1.6414 |
| FEA (754) | 1.5827 | 0.0713 | 1.5942 | 1.6405 | 0.0373 | 1.6439 |
| ADH (501) | 1.6261 | 0.0613 | 1.6342 | 1.6343 | 0.0323 | 1.6385 |
| DCIS (730) | 1.6433 | 0.0674 | 1.6509 | 1.6308 | 0.0441 | 1.6357 |
| IC (533) | 1.6312 | 0.0844 | 1.6426 | 1.6131 | 0.0596 | 1.6212 |

**Table S2.** Paired null model comparison results for the Minkowski dimension (*Dm*) across seven breast tissue subtypes, ordered by clinical progression sequence. For each subtype, the real median *Dm* is compared against the median *Dm* of a geometry-matched smooth ellipse null model, constructed by perturbing the semi-major axis, semi-minor axis, and orientation of each segmented nucleus with small Gaussian noise (5 realisations per image). Statistical significance was assessed using the paired Wilcoxon signed-rank test; p-values are reported in scientific notation. Effect size is reported as Cohen's *d* for paired differences, computed as the mean with Bessel's correction. N pairs denote the number of valid paired real–null observations per subtype.

| Subtype | N pairs | ***Dm* Median (Real)** | ***Dm* Median (Null)** | ***p*-value** | **Cohen's *d*** |
|---|---|---|---|---|---|
| N | 476 | 1.3322 | 1.3057 | 7.6691e-69 | 1.392 |
| PB | 775 | 1.3229 | 1.2969 | 1.6723e-121 | 1.475 |
| UDH | 507 | 1.3504 | 1.3224 | 2.2970e-78 | 0.925 |
| FEA | 754 | 1.3299 | 1.3016 | 8.7255e-107 | 1.096 |
| ADH | 501 | 1.3137 | 1.2915 | 5.7407e-75 | 1.012 |
| DCIS | 730 | 1.2833 | 1.2650 | 1.6361e-113 | 0.832 |
| IC | 533 | 1.2691 | 1.2538 | 6.2198e-66 | 0.423 |

**Table S3.** Paired null model comparison results for the Correlation dimension (*Dc*) across seven breast tissue subtypes, ordered by clinical progression sequence. For each subtype, the real median *Dc* is compared against the median *Dc* of two null spatial processes: Complete Spatial Randomness (CSR; Poisson process) and a Thomas cluster process (clustered null). Both null models were conditioned on the observed centroid count and domain dimensions of each image (20 realisations per image), ensuring paired comparisons. Statistical significance was assessed using the paired Wilcoxon signed-rank test; p-values are reported in scientific notation. N pairs denote the number of valid paired real–null observations per subtype.

| Subtype | N pairs | *Dc* Median (Real) | CSR Median | *p* (vs. CSR) | Clustered Null Median | *p* (vs. Clustered) |
|---|---|---|---|---|---|---|
| N | 476 | 1.7985 | 1.8531 | 3.1970e-51 | 1.7733 | 2.6371e-12 |
| PB | 775 | 1.7424 | 1.8491 | 8.1004e-110 | 1.7703 | 5.1954e-08 |
| UDH | 507 | 1.7867 | 1.8465 | 4.6655e-47 | 1.7042 | 1.3261e-26 |
| FEA | 754 | 1.6881 | 1.8425 | 3.9559e-115 | 1.6172 | 8.0295e-23 |
| ADH | 501 | 1.8047 | 1.8432 | 2.2019e-35 | 1.6922 | 3.3231e-46 |
| DCIS | 730 | 1.8059 | 1.8450 | 1.7693e-58 | 1.7745 | 5.0369e-15 |
| IC | 533 | 1.8118 | 1.8457 | 1.6793e-51 | 1.8257 | 2.8496e-01 |
